# Interleukin-1 receptor antagonist gene (*IL1RN*) variants modulate the cytokine release syndrome and mortality of SARS-CoV-2

**DOI:** 10.1101/2023.01.09.23284348

**Authors:** Mukundan Attur, Christopher Petrilli, Samrachana Adhikari, Eduardo Iturrate, Xiyue Li, Stephanie Tuminello, Nan Hu, Aravinda Chakravarti, David Beck, Steven B. Abramson

**Affiliations:** Division of Rheumatology, Department of Medicine, NYU Langone Orthopedic Hospital, NYU Langone Health, NY -10003; Department of Medicine, NYU Grossman School of Medicine, NYU Langone Health, New York, NY -10016; Division of Biostatistics, Department of Population Health, NYU Grossman School of Medicine, New York, NY-10016; Center for Human Genetics and Genomics, NYU Grossman School of Medicine, New York, NY-10016

**Keywords:** SARS-CoV-2, inflammation, *IL-1RN*, genetic variation and mortality

## Abstract

**Objective:** To explore the regulation of the inflammatory response in acute SARS-CoV-2 infection, we examined effects of single nucleotide variants (SNVs) of *IL1RN*, the gene encoding the anti-inflammatory IL-1 receptor antagonist (IL-1Ra), on the cytokine release syndrome and mortality.

**Methods:** We studied 2589 patients hospitalized with SARS-CoV-2 between March 2020 and March 2021 at NYU Langone’s Tisch Hospital. CTA and TTG haplotypes formed from three SNVs (rs419598, rs315952, rs9005) and the individual SNVs of the *IL1RN* gene were assessed for association with laboratory markers of the cytokine release syndrome (CRS) and mortality.

**Results:** Mortality in the population was 15.3%, and was lower in women than men (13.1% vs.17.3%, p<0.0003). Carriers of the CTA-1/2 *IL1RN* haplotypes exhibited *decreased* inflammatory markers and *increased* plasma IL-1Ra relative to TTG carriers. Decreased mortality among CTA-1/2 carriers was observed in male patients between the ages of 55-74 [9.2% vs. 17.9%, p=0.001]. Evaluation of individual SNVs of the *IL1RN* gene (rs419598, rs315952, rs9005) indicated that carriers of the *IL1RN* rs419598 CC SNV exhibited lower inflammatory biomarker levels, and was associated with reduced mortality compared to the CT/TT genotype in men (OR 0.49 (0.23 – 1.00); 0.052), with the most pronounced effect observed between the ages of 55-74 [5.5% vs. 18.4%, p<0.001].

**Conclusion:** The *IL1RN* haplotype CTA, and sequence variant of rs419598 are associated with attenuation of the cytokine release syndrome and decreased mortality in males with acute SARS-CoV2 infection. The data suggest that *IL1RN* modulates the COVID-19 cytokine release syndrome via endogenous “ anti-inflammatory” mechanisms.

**Significance statement:** We provide evidence that variants of *IL1RN* modulate the severity of SARS-CoV-2 infection. The *IL1RN CTA haplotype and* rs419598 CC single nucleotide variant are associated with decreased plasma levels of inflammatory markers, interleukin-1 beta (IL-1β), interleukin-6 (IL-6), interleukin-2 (IL-2), C-reactive protein (CRP), D-dimer, ferritin, and procalcitonin, in association with higher levels of IL-1Ra and IL-10, anti-inflammatory proteins. Both haplotype CTA and rs419598 CC genotype are associated with a significant reduction in the mortality of men. These data provide genetic evidence that inflammasome activation and the IL-1 pathway plays an important role in the mortality and morbidity associated with severe SARS-CoV-2 infection, and that genetic regulation of inflammatory pathways by variants of *IL1RN* merits further evaluation in severe SARS-CoV-2 infection.

## Introduction

SARS-CoV-2 infections may be asymptomatic or cause only mild symptoms in most cases. However, acute respiratory distress syndrome (ARDS), multi-organ failure, and death occur in nearly 10–20% of cases, especially in the elderly or those with pre-existing comorbidities (1, 2). Patients with severe SARS-CoV-2 symptoms are characterized by hyper-inflammatory responses with unusually high serum cytokine levels similar to those observed in the cytokine release syndrome (CRS), including, though not limited to, elevations of IL-1β, IL-2 and IL-6 (3-5). Clinical evidence that systemic cytokine release contributes to poor outcomes in patients with SARS-CoV-2 is supported by the clinical effectiveness of targeted anti-inflammatory agents, including corticosteroids, Janus kinase (JAK) inhibitors, IL-6 inhibitors, and anakinra a recombinant human interleukin-1 receptor antagonist (IL-1Ra), recently approved by the FDA for the treatment of hospitalized patients with SARS-CoV-2 (6-10).

The dysregulation of immune mechanisms leading to the hyper-inflammation of severe SARS-CoV-2 is poorly understood. In a recent study, Sefik and colleagues presented data to suggest that inflammasome activation in SARS-CoV-2-infected macrophages causing the release of IL-1 was a key driver of COVID-19 pathology (11, 12). An algorithm that analyzed the cytokines and clinical variables contributing to the CRS indicated that the anti-inflammatory cytokines IL-10 and IL-1Ra were significantly more elevated early in severe COVID-19 disease than the more frequently characterized cytokines such as IL-6, IL-1β, and tumor necrosis factor (TNFα) (11, 13). IL-1Ra, an endogenous anti-inflammatory protein, competitively binds to the IL-1 receptor and modulates the production of cytokines such as IL-1, IL-6, and TNF-α. The *IL1RN* gene regulates IL-1Ra production; several single nucleotide variants (SNVs) in this gene have been associated with multiple inflammatory syndromes, including systemic lupus erythematosus, inflammatory bowel disease, obesity, and insulin resistance (14-18).

We have previously shown that specific *IL1RN* haplotypes (TTG, CTA) comprising of the three variants rs419598, rs315952, and rs9005 are associated with inflammation and disease severity of osteoarthritis and rheumatoid arthritis (RA) (19). Notably, RA patients carrying the *IL1RN* CTA haplotype exhibit lower disease activity scores (DAS), plasma CRP, and IL-6 in association with increased IL-1Ra (19).

Building on that observation, this study sought to determine whether the same *IL1RN* polymorphisms (SNVs rs419598, rs315952, and rs9005) are similarly associated with inflammation and mortality in patients with moderate to severe SARS-CoV-2. We performed low coverage whole-genome sequencing followed by imputation on 2,589 patients admitted to NYU Langone’s Tisch Hospital from March 1, 2020, through March 1, 2021. We analyzed the association of these *IL1RN* genotypes with inflammatory markers of the CRS and mortality.

## Results

### Patient Population Demographics and Clinical Characteristics

We obtained laboratory and health information as part of standard clinical care from 2,589 hospitalized COVID-19 PCR positive patients admitted to NYU Langone’s Tisch Hospital between March, 1 2020 through March 1, 2021. The alpha and beta SARS-CoV-2 variants were the most prevalent during this time. Low coverage (1.2X) whole-genome sequencing data followed by imputation were obtained on all 2,589 patients, from which *IL1RN* genotype data were obtained, as described elsewhere (20). The mean (standard deviation) age in years and BMI for the cohort were 61.2 (18.7) (range 19 - 102) years and 30.43 (7.71), respectively. Males represented 53.3% of the sample. *IL1RN* rs419598, rs315952, and rs9005 genotype data were available for all patients. Biomarkers noted in the clinical EHR for IL-1β, IL-2, and IL-6 were available for 642, 645, and 1229 subjects, respectively, whereas other clinical inflammation markers were available for more than 2000 subjects (Supplemental Table 1).

### Biochemical and Inflammatory Markers Associated with Mortality

We first determined which inflammatory markers were associated with an increased likelihood of mortality in these hospitalized SARS-CoV-2 patients. There were 397 (15.3%) deaths among the 2589 patients. As expected, age, male gender, and BMI were associated with increased mortality (Table 1). Consistent with prior literature (6, 21), those who died exhibited significantly higher baseline or maximum levels of markers associated with the cytokine release syndrome (IL-6, IL-1β, CRP, procalcitonin, ferritin, D-dimer) (Table 1). Table 1 also shows that mortality was associated with reduced levels of complement components C3 and C4.

**Table 1:**
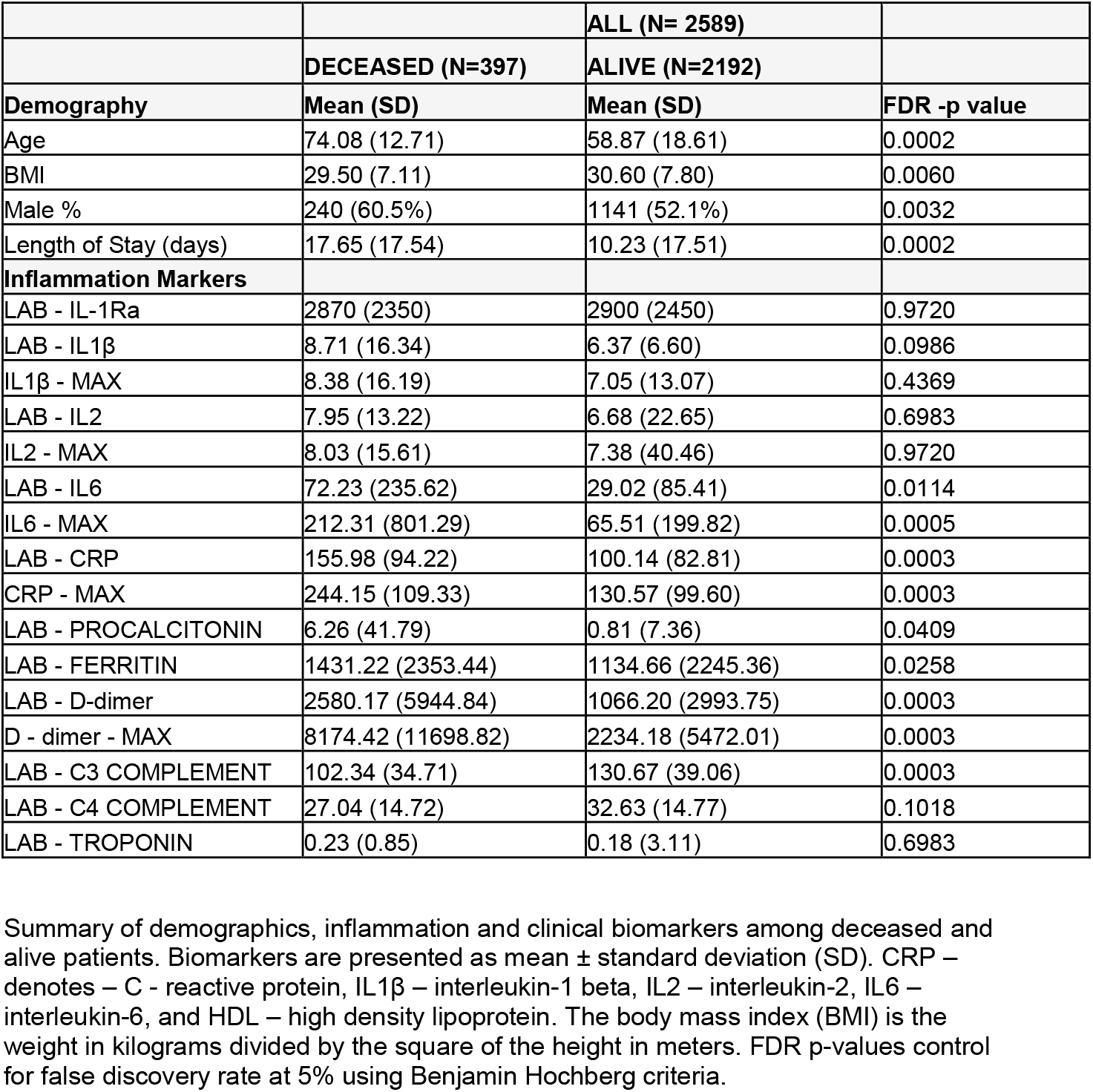
Levels of inflammation biomarkers associated with COVID-19 mortality.

|  |  | ALL (N= 2589) |  |
| --- | --- | --- | --- |
|  | DECEASED (N=397) | ALIVE (N=2192) |  |
| Demography | Mean (SD) | Mean (SD) | FDR -p value |
| Age | 74.08 (12.71) | 58.87 (18.61) | 0.0002 |
| BMI | 29.50 (7.11) | 30.60 (7.80) | 0.0060 |
| Male % | 240 (60.5%) | 1141 (52.1%) | 0.0032 |
| Length of Stay (days) | 17.65 (17.54) | 10.23 (17.51) | 0.0002 |
| Inflammation Markers |  |  |  |
| LAB - IL-1Ra | 2870 (2350) | 2900 (2450) | 0.9720 |
| LAB - IL1β | 8.71 (16.34) | 6.37 (6.60) | 0.0986 |
| IL1β - MAX | 8.38 (16.19) | 7.05 (13.07) | 0.4369 |
| LAB - IL2 | 7.95 (13.22) | 6.68 (22.65) | 0.6983 |
| IL2 - MAX | 8.03 (15.61) | 7.38 (40.46) | 0.9720 |
| LAB - IL6 | 72.23 (235.62) | 29.02 (85.41) | 0.0114 |
| IL6 - MAX | 212.31 (801.29) | 65.51 (199.82) | 0.0005 |
| LAB - CRP | 155.98 (94.22) | 100.14 (82.81) | 0.0003 |
| CRP - MAX | 244.15 (109.33) | 130.57 (99.60) | 0.0003 |
| LAB - PROCALCITONIN | 6.26 (41.79) | 0.81 (7.36) | 0.0409 |
| LAB - FERRITIN | 1431.22 (2353.44) | 1134.66 (2245.36) | 0.0258 |
| LAB - D-dimer | 2580.17 (5944.84) | 1066.20 (2993.75) | 0.0003 |
| D - dimer - MAX | 8174.42 (11698.82) | 2234.18 (5472.01) | 0.0003 |
| LAB - C3 COMPLEMENT | 102.34 (34.71) | 130.67 (39.06) | 0.0003 |
| LAB - C4 COMPLEMENT | 27.04 (14.72) | 32.63 (14.77) | 0.1018 |
| LAB - TROPONIN | 0.23 (0.85) | 0.18 (3.11) | 0.6983 |

Summary of demographics, inflammation and clinical biomarkers among deceased and alive patients. Biomarkers are presented as mean ± standard deviation (SD). CRP – denotes – C - reactive protein, IL1β – interleukin-1 beta, IL2 – interleukin-2, IL6 – interleukin-6, and HDL – high density lipoprotein. The body mass index (BMI) is the weight in kilograms divided by the square of the height in meters. FDR p-values control for false discovery rate at 5% using Benjamin Hochberg criteria.

Since a number of cytokines of interest were not measured in the course of patient care reported in the EHR, we analyzed 359 additional plasma (all available samples from NYU Center for Biospecimen Research and Development biobank) from the 2,589 patient cohort using a multiplex ELISA assay, and compared to age, sex and BMI matched healthy controls (n=22). The results show elevations of additional cytokines not reported in the EHR, indicative of the CRS in these patients, including IL-1β, IL-1α, IL-5, IL-6, IL-8, IL-17, TNFα, VEGF and Interferon (IFN) *α* (Figure 1 Panel A). As shown in Fig. 1 Panel A a number of these cytokines were strikingly elevated, exceeding 10-fold normal levels, including IL-10, IL-8, IL-1Ra and IL-6. Of note, two of these four most elevated cytokines were IL-1Ra (12-fold) and IL-10 (62-fold), “ anti-inflammatory” proteins. Correlation analysis indicated moderate positive and significant correlations (r=0.19 - 0.5) between elevated IFN-*α* and MCP-1(CCL2), IL-10, IL-1Ra, IL-6 and TNFα. (Figure Panel B).

**Figure 1:**
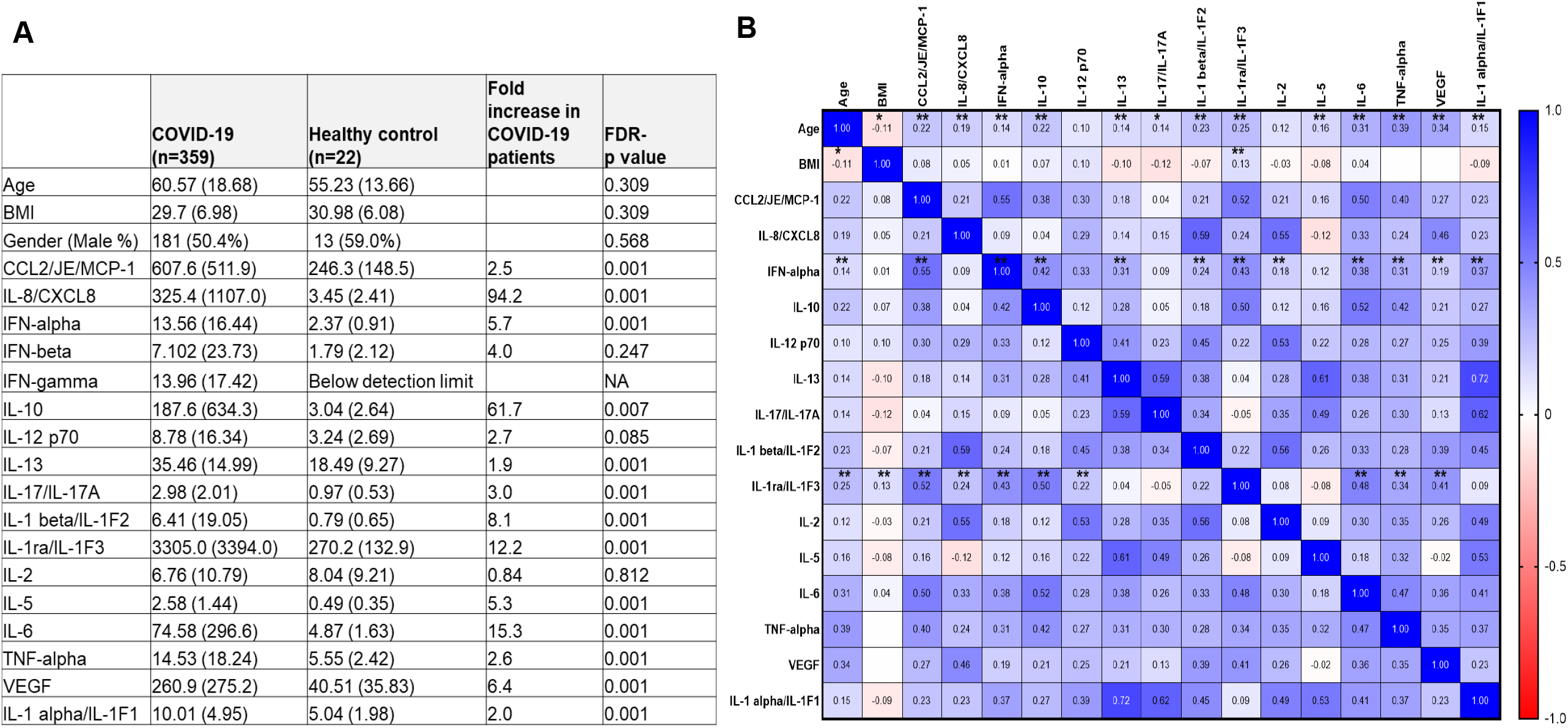
Elevated levels interferon and cytokines and chemokines in SARS-CoV-2 patients. **1A**. Plasma samples of sub-set of acute COVID-19 patients (N=371) and healthy controls (n=22) were screened to measure the levels of multiple cytokines: CCL2, IL-8, IFNα, IFN-β, IFN-γ, IL-10, IL-12p70, IL-13, IL-17A, IL-1β, IL-1RA, IL-2, IL-5, IL-6, TNF-α, VEGF, and IL-1α. A customized version of the Human XL Cytokine Premixed Kit (#FCSTM18, R&D Systems) was used to simultaneously measure the 17 analytes through a magnetic bead–based fluorescence sandwich ELISA. Cytokine biomarkers are presented as mean ± standard deviation (SD). Adjusted p-values control for false discovery rate at 5% using Benjamin Hochberg criteria. **1B:** Spearman correlation of multiple cytokines. chemokines and interferon. The * indicates p=0.05 and **p=0.001.

### *IL1RN* CTA haplotype is associated with decreased inflammation and mortality

Based upon our prior studies of *IL1RN* polymorphisms in rheumatoid and osteoarthritis, we compared carriers of *IL1RN* CTA-1/2 versus TTG haplotypes (TTG-0/1/2) for association with inflammatory markers in acute SARS-CoV-2 patients. Haplotypes CTA and TTG are two of the nine potential haplotypes that can be constructed from these three SNPs that have a frequency of >1%, and are found on the same locus. The carriers of the CTA-1/2 haplotypes exhibited significantly lower levels of inflammatory markers (IL-1*β*, IL-2, IL-6, D-dimer) and *higher* levels of IL-1Ra relative to TTG-0/1/2 and CTA-1/TTG-1 heterozygote haplotype carriers (Table 2). We also observed decreased age, sex and BMI adjusted mortality among hospitalized CTA carriers relative to TTG carriers that did not reach statistical significance in the total cohort. However, sex differences in the association were noted; mortality was decreased in male carriers of the CTA-1/2 haplotype (11.3% vs. 17.9%; unadjusted p = 0.06, age and BMI adjusted OR 0.73 (0.41 - 1.24); p= 0.271) (Table 2), which was not observed in female patients. The decreased mortality among men conferred by the CTA haplotype was best appreciated when mortality was assessed according to age. Specifically, decreased mortality among CTA-1/2 carriers was significantly reduced in male patients between the ages of 55-74 [9.2% vs. 17.9%, p=0.001] (Supplemental Figure 1).

**Table 2:** Comparison of *IL1RN* haplotype CTA-1/2 with TTG-0/1/2 and CTA-1/TTG-1 heterozygotes.

|  | *IL1RN* ALL patients (N=2589) |  |  |  |  |  |
| --- | --- | --- | --- | --- | --- | --- |
|  | CTA-1/2<br>(N=263) | CTA-0 (TTG-1/2)<br>(N=1918) | CTA-1/TTG-1<br>(N=408) | CTA-1/2 Vs. TTG-0/1/2 | TTG-0/1/2 vs.CTA1/TTG1 | CTA-1/2 vs. CTA-1/TTG-1 |
| **Demography** | **Mean (SD)** | **Mean (SD)** | **Mean (SD)** | **FDR - p value** | **FDR - p value** | **FDR - p value** |
| Age | 58.44 (18.25) | 61.37 (18.58) | 62.16 (19.14) | 0.0453 | 0.8055 | 0.0351 |
| BMI | 30.48 (6.82) | 30.44 (7.48) | 30.40 (9.18) | 0.9193 | 0.9438 | 0.8969 |
| Gender (Male %) | 150 (57.0%) | 1008 (52.6%) | 222 (54.4%) | 0.2955 | 0.8055 | 0.8355 |
| **Inflammation Markers** |  |  |  |  |  |  |
| LAB - IL-1Ra | 3910 (3630) | 2700 (2040) | 3260 (3140) | 0.0458 | 0.4238 | 0.5001 |
| LAB - IL1β | 5.85 (3.02) | 7.32 (11.43) | 6.44 (5.19) | 0.0436 | 0.5111 | 0.5001 |
| LAB - IL2 | 5.10 (2.28) | 7.47 (23.03) | 5.63 (6.02) | 0.0458 | 0.4238 | 0.5001 |
| LAB - IL6 | 27.56 (55.97) | 42.91 (179.57) | 37.91 (103.79) | 0.0623 | 0.6544 | 0.5001 |
| IL1β - MAX | 5.89 (2.72) | 7.89 (15.74) | 6.37 (4.62) | 0.0302 | 0.4238 | 0.5001 |
| IL2 - MAX | 4.63 (2.39) | 7.98 (39.71) | 6.10 (12.47) | 0.0828 | 0.6241 | 0.5001 |
| IL6 - MAX | 84.24 (276.51) | 111.86 (455.03) | 106.04 (337.35) | 0.3443 | 0.8440 | 0.5824 |
| LAB - CRP | 103.79 (75.66) | 111.66 (88.14) | 103.86 (87.96) | 0.1918 | 0.4238 | 0.9916 |
| CRP - MAX | 136.20 (99.39) | 152.72 (110.11) | 147.97 (116.04) | 0.0458 | 0.6241 | 0.5001 |
| LAB - PROCALCITONIN | 0.45 (0.91) | 1.98 (20.10) | 0.81 (3.25) | 0.0186 | 0.4238 | 0.4524 |
| LAB - FERRITIN | 961.24 (1189.92) | 1272.14 (2662.63) | 1166.16 (2710.13) | 0.0186 | 0.6241 | 0.5001 |
| LAB - D-dimer | 933.97 (1825.80) | 1367.67 (3814.58) | 1185.69 (3639.27) | 0.0237 | 0.6241 | 0.5001 |
| D-dimer - MAX | 2052.95 (3430.96) | 3476.49 (7614.07) | 3131.53 (7332.19) | 0.0013 | 0.6241 | 0.3094 |
| Mortality | 31 (11.8%) | 298 (15.5%) | 68 (16.6%) | 0.83 (0.54 - 1.26); 0.401 | 0.99 (0.73 - 1.35); 0.934 | 0.81 (0.49 - 1.31); 0.392 |
| Mortality (Men only) | 17 (11.3%) | 180 (17.9%) | 42 (18.9%) | 0.73 (0.41 - 1.24); 0.271 | 0.98 (0.67 - 1.47); 0.924 | 0.72 (0.38 - 1.35); 0.322 |
| Mortality (women only) | 14 (12.4%) | 118 (13.0%) | 26 (14.0%) | 1.00 (0.50 - 1.87); 0.996 | 1.01 (0.62 - 1.68); 0.966 | 1.03 (0.47 - 2.21); 0.942 |

Summary of demographics and inflammation biomarkers by haplotypes. Biomarkers are presented as mean ± standard deviation (SD). CRP – denotes – C - reactive protein, IL1β – interleukin-1 beta, IL6 – interleukin-6. The body mass index (BMI) is the weight in kilograms divided by the square of the height in meters. FDR p-values control for false discovery rate at 5% using Benjamin Hochberg criteria within each subgroup of biomarkers. For mortality comparison between haplotypes - odds ratio [aOR (95% Confidence intervals)] and p values derived from multivariable logistic regression adjusting for age, gender and BMI; and for age and BMI for gender specific mortality.

### The *IL1RN* rs419598 CC genotype is associated with lower plasma inflammatory biomarkers

Having shown that the CTA-1/2 haplotype is associated with decreased inflammatory biomarkers and mortality (in men) as compared to TTG-0/1/2, we next evaluated each single nucleotide variant of the *IL1RN* gene (rs419598, rs315952 and rs9005) individually. As shown in Table 3, multiple biomarkers of inflammation in COVID-19 were significantly lower in patients with the *IL1RN* rs419598 CC genotype compared to those with CT or TT. In particular, the CC genotype was associated with significantly lower plasma levels of IL-1β, IL-2, IL-6, ferritin, procalcitonin, CRP, and D-dimer. The differences between genotypes were striking: in particular, maximum levels of IL-1β (p<0.001), IL-2 (p = 0.048), IL-6 (p=0.030), and D-dimer (p<0.001) were 1.4- to 2-fold higher in CT/TT genotypes (Table 3). Conversely, similar to the CTA haplotype findings, and in contrast to *decreased* inflammatory cytokine levels observed in *IL1RN* rs419598 CC individuals, these patients exhibited numerically higher plasma IL-1Ra levels, the gene product of *IL1RN* and an endogenous *anti-inflammatory* cytokine (Table 3). The *IL1RN* rs419598 CC vs. CT/TT genotype-dependent decrease of inflammation-associated biomarkers was comparable in both males and females, except for IL-6 - MAX and CRP, for which no significant differences between genotypes were observed in women (Table 4). An additional finding of interest was that mean plasma IL-1Ra levels exceeded 4950 pg/ml, two-fold higher in women carriers of the CC compared to carriers with the CT/TT genotype (Table 4).

**Table 3.** Patients with the *IL1RN* rs419598 CC genotype exhibit lower plasma inflammation cytokine levels and mortality.

|  | *IL1RN* - rs419598 |  |  |
| --- | --- | --- | --- |
|  | CC (N=175) | CT/TT (N=2414) |  |
| **Demography** | **Mean (SD)** | **Mean (SD)** | **FDR -p value** |
| Age | 60.5(19.5) | 61.3(18.6) | 0.619 |
| BMI | 30.2(6.84) | 30.5(7.77) | 0.659 |
| Male % | 90 (51.4%) | 1290 (53.4%) | 0.659 |
| **Inflammation Markers** |  |  |  |
| LAB - IL-1Ra | 3680(3720) | 2840(2320) | 0.199 |
| LAB - IL1β | 5.55(1.93) | 7.13(10.4) | 0.002 |
| LAB - IL2 | 5.04(1.92) | 7.06(20.6) | 0.024 |
| LAB - IL6 | 24.1(35.7) | 41.7(166) | 0.006 |
| IL1β - MAX | 5.45(0.853) | 7.57(14.1) | < 0.001 |
| IL2 - MAX | 4.58(1.90) | 7.51(35.6) | 0.048 |
| IL6 - MAX | 58.9(178) | 112(435) | 0.030 |
| LAB - CRP | 96.4(67.9) | 111(88.1) | 0.026 |
| CRP - MAX | 132(95.4) | 152(111) | 0.026 |
| LAB - PROCALCITONIN | 0.349(0.862) | 1.74(18.0) | 0.001 |
| LAB - FERRITIN | 897(1110) | 1250(2630) | 0.002 |
| LAB - D-dimer | 896(1780) | 1320(3730) | 0.015 |
| D-dimer - MAX | 1910(3230) | 3380(7460) | < 0.001 |
| Mortality % | 20 (11.4%) | 377 (15.6%) | 0.66 (0.39 – 1.09); 0.12 |

Summary of demographics and inflammation biomarkers by genotype. Biomarkers are presented as mean ± standard deviation (SD). CRP – denotes – C - reactive protein, IL1β – interleukin-1 beta, IL2 – interleukin -2, IL6 – interleukin-6. The body mass index (BMI) is the weight in kilograms divided by the square of the height in meters. FDR p-values control for false discovery rate at 5% using Benjamin Hochberg criteria within each subgroup of biomarkers. For mortality comparison between genotype- odds ratio [aOR (95% Confidence intervals)] and p values derived from multivariable logistic regression adjusting for age, gender and BMI.

**Table 4:** Association of *IL1RN* rs419598 genotype with inflammation markers, and mortality (men vs. women)

|  | Men (N=1380) |  |  | Women (N=1208) |  |  |
| --- | --- | --- | --- | --- | --- | --- |
|  | CC (N=90) | CT/TT (1290) | FDR –p-value | CC (N=85) | CT/TT (N=1123) | FDR –p-value |
| **Demography** |  |  |  |  |  |  |
| Age | 62.0(17.7) | 62.5(15.9) | 0.8190 | 58.9(21.3) | 59.8(21.2) | 0.6810 |
| BMI | 29.0(5.56) | 30.0(6.67) | 0.1760 | 31.6(7.80) | 30.9(8.84) | 0.6810 |
| **Inflammation Markers** |  |  |  |  |  |  |
| LAB - IL-1Ra | 2410(1600) | 2930(2340) | 0.2280 | 4950(4750) | 2730(2290) | 0.1069 |
| LAB - IL1β | 5.64(2.32) | 6.59(8.45) | 0.1501 | 5.40(0.949) | 8.05(13.0) | 0.0260 |
| LAB - IL2 | 5.16(1.88) | 7.18(24.0) | 0.1501 | 4.83(2.01) | 6.85(12.8) | 0.0854 |
| LAB - IL6 | 25.9(41.4) | 43.5(168) | 0.0748 | 21.6(25.9) | 38.8(164) | 0.0910 |
| IL1β - MAX | 5.37(0.765) | 7.30(14.4) | 0.0238 | 5.59(1.00) | 8.04(13.6) | 0.0476 |
| IL2 - MAX | 4.64(1.74) | 7.78(43.2) | 0.1744 | 4.47(2.21) | 7.04(14.8) | 0.0628 |
| IL6 - MAX | 46.8(84.1) | 136(529) | 0.0003 | 74.9(256) | 74.0(215) | 0.9830 |
| LAB - CRP | 98.6(65.3) | 118(89.6) | 0.0316 | 93.5(71.8) | 99.4(84.6) | 0.7319 |
| CRP - MAX | 133(88.8) | 162(111) | 0.0234 | 129(105) | 136(109) | 0.7800 |
| LAB - PROCALCITONIN | 0.244(0.525) | 1.07(6.11) | 0.0003 | 0.490(1.16) | 2.75(27.5) | 0.0628 |
| LAB - FERRITIN | 1240(1330) | 1500(2730) | 0.1500 | 442(374) | 871(2450) | 0.0013 |
| LAB - D-dimer | 715 (1380) | 1410 (4320) | 0.0003 | 1140(2190) | 1200(2670) | 0.9078 |
| D-dimer - MAX | 1970(3590) | 3760 (7890) | 0.0003 | 1830(2690) | 2820(6770) | 0.0628 |
| Mortality % | 9 (10.0%) | 230 (17.8%) | 0.49 (0.23 – 1.00);0.052 | 11 (12.9%) | 147 (13.1%) | 1.0 (0.48 – 2.09) 0.99 |

Summary of demographics and inflammation biomarkers by genotype. *IL1RN* – interleukin-1 receptor antagonist gene, CRP – C - reactive protein, IL1β – interleukin-1 beta, IL2 – interleukin-2, IL6 –interleukin-6. The body mass index (BMI) is the weight in kilograms divided by the square of the height in meters. Biomarkers data are presented as mean ± standard deviation (SD). Adjusted p-values control for false discovery rate at 5% using Benjamin Hochberg criteria within each subgroup of biomarkers. For mortality comparison between genotype - odds ratio [aOR (95% Confidence intervals)] and p values derived from multivariable logistic regression adjusting for age and BMI.

We also examined the association of two other *IL1RN* SNVs (rs315952, rs9005), part of the *IL1RN* haplotype of interest (19). Patients with the *IL1RN* rs315952 CC genotype exhibited lower levels of IL1β and IL-2 (Supplemental Table 2), but did not affect other inflammatory markers. The SNV rs9005 AA genotypes, in close linkage disequilibrium (LD) to rs419598 (D’/r2 0.81/0.61), was associated with elevations of IL-1Ra and decreased levels of IL-1β, IL-2, and D-dimer Max (Supplemental Table 3).

### The *IL1RN* CC rs419598 SNV is associated with decreased SARS-CoV-2 mortality in men

As noted above, male carriers of the CTA-1/2 haplotype exhibited decreased mortality compared to carriers of the TTG-0/1/2 haplotype, an association observed in men but not women (Table 2). We therefore next determined whether the lower levels of mortality-associated biomarkers in rs419598 CC patients were associated with decreased mortality Table 4 compares the association of *IL1RN* rs419598 genotype with inflammation and mortality in men and women. As was the case with the CTA haplotype noted above, the CC genotype was associated with decreased mortality in men [17.8% vs. 10%, (OR = 0.49 (0.23 – 1.00); p=0.052] in association with lower levels of inflammatory biomarkers (IL-6, IL-1β, IL-2, CRP, procalcitonin, D-dimer). The *IL1RN* rs419598 genotype conferred no such mortality benefit in women, although women carriers of the CC genotype exhibited significantly lower levels of IL-1β, ferritin and procalcitonin, as well as numerically lower levels of IL-2, IL-6 and D-dimer. Of note, plasma levels of IL-1Ra were strikingly high in women carriers of the CC genotype (4950.0 vs. 2730.0; p=0.1069). In separate analyses, the mortality rate did not differ between genotypes for men or women across two other *IL1RN* SNVs (rs315952 and rs9005) studied (Supplemental Tables 2 & 3).

To further explore differences in genetic associations with mortality between males and females we examined inflammation markers and mortality according to sex (Table 5). Women patients were younger and had higher BMI (Table 5). Baseline and maximal elevations of IL-1β and IL-2 were comparable between men and women, while IL-6 Max was lower in women as has been reported (Table 5) (5). Plasma levels of the inflammatory markers ferritin, CRP and D-dimer Max were also lower in women compared to men.

**Table 5.** Inflammation markers and mortality according to gender.

|  | **COVID-19 Patients (N= 2589)** |  |  |
| --- | --- | --- | --- |
|  | **Men (N=1380)** | **Women (N=1208)** |  |
| **Demography** | **Mean (SD)** | **Mean (SD)** | **FDR -p value** |
| Age | 62.45 (16.00) | 59.78 (21.20) | < 0.001 |
| BMI | 29.95 (6.60) | 30.99 (8.77) | < 0.001 |
| **Inflammation Markers** |  |  |  |
| LAB - IL-1Ra | 2696.65 (1805.49) | 2697.45 (2735.4) | 0.9900 |
| LAB - IL1β | 6.52 (8.18) | 7.87 (12.59) | 0.2619 |
| LAB - IL2 | 7.04 (23.13) | 6.71 (12.36) | 0.8808 |
| LAB - IL6 | 42.39 (163.23) | 37.64 (157.91) | 0.7969 |
| IL1β - MAX | 7.17 (13.92) | 7.87 (13.13) | 0.7583 |
| IL2 - MAX | 7.57 (41.74) | 6.86 (14.28) | 0.8808 |
| IL6 - MAX | 130.06 (512.83) | 74.04 (217.57) | 0.0260 |
| LAB - CRP | 117.05 (88.41) | 98.97 (83.81) | 0.0043 |
| CRP - MAX | 160.71 (109.87) | 135.33 (108.82) | 0.0043 |
| LAB - PROCALCITONIN | 1.02 (5.92) | 2.60 (26.54) | 0.2232 |
| LAB - FERRITIN | 1487.98 (2660.92) | 841.82 (2364.73) | 0.0013 |
| LAB - D-dimer | 1367.10 (4199.01) | 1194.22 (2635.99) | 0.4111 |
| D-dimer - MAX | 3650.46 (7703.49) | 2757.00 (6583.86) | 0.0163 |
| Mortality | 239 (17.3%) | 158 (13.1%) | 1.55 (1.22 - 1.96); 0.0003 |

Laboratory inflammatory biomarkers of patients hospitalized with SARS-Cov-2 infected COVID-19 patients by gender. Biomarkers data shown are presented as mean ± standard deviation (SD). CRP – denotes – C - reactive protein, IL1β – interleukin-1 beta, IL6 – interleukin-6, Baseline and Max values – Initial and maximum value biomarkers – when multiple values are available. The body mass index (BMI) is the weight in kilograms divided by the square of the height in meters. FDR p-values control for false discovery rate at 5% using Benjamin Hochberg criteria within each subgroup of biomarkers. For mortality comparison - odds ratio [aOR (95% Confidence intervals)] and p values derived from multivariable logistic regression adjusting for age and BMI.

### Outcomes According to Age

As shown in Figure 2A, and as expected, SARS-CoV-2 mortality rate increased with age, and was greater in men than in women [OR 1.55 (95% CI 1.22 - 1.96); 0.0003]. Consistent with the *IL1RN* data referenced above (Table 4), Figure 2B demonstrates that the rs419598 CC SNV is associated with decreased mortality in men shown as a continuous variable (adjusted OR 0.38 (95% CI 0.15-0.96, p<0.04). Figures 2C and 2D examine mortality in men according to decade. As shown, for cohorts between the ages of 55-74, the SNV CC rs 4198598 is associated with significantly decreased mortality (5.5% vs. 18.4%, p<0.001). These data are similar to those noted above, showing decreased mortality in male CTA-1/2 carriers between the ages of 55-74 [9.2% vs. 18.0%, p=0.001] (Supplemental Figure 1).

**FIGURE 2:**
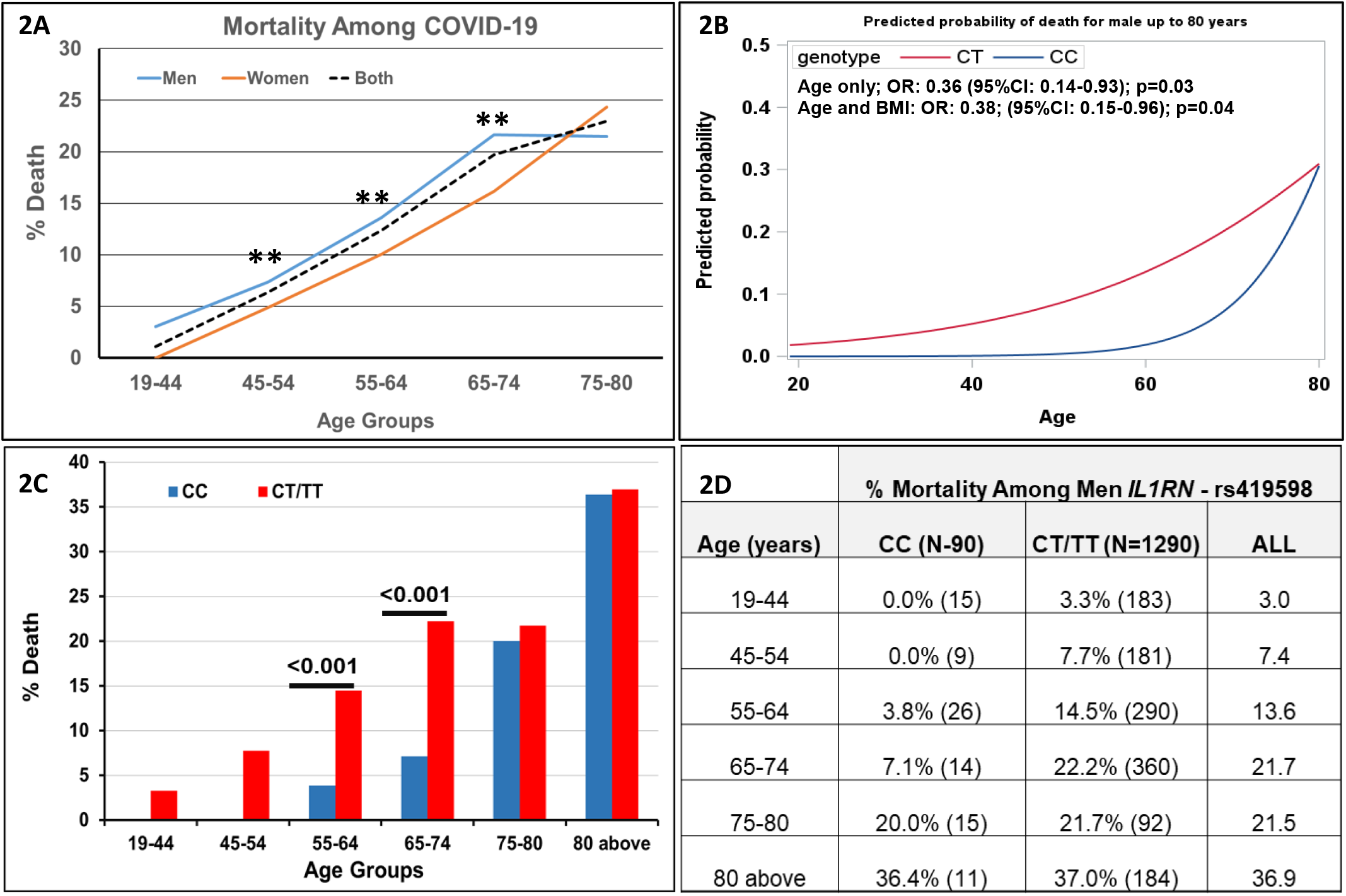
*IL1RN* rs419598 Protective Genotype is Associated with Decreased Mortality Among Men. **2A**. Mortality rate by age categories overall and by sex among SARS-CoV-2 infected COVID-19 patients. **2B**. Model predicted mortality rate for men by *IL1RN* rs419598 genotype. Unadjusted odds ratios (OR) and 95% confidence interval were estimated using logistic regression adjusted for age or age and BMI. **2C**. Mortality rate among men by *IL1RN* rs419598 (CC vs. CT/TT) genotype by different age groups **2D**. Mortality as percentage by decade is shown for *IL1RN* rs419598 CC and CT/TT variant carriers among men, while total number of patient are shown in parentheses for each age group. *IL1RN* – denotes interleukin-1 receptor antagonist gene

Figures 2B and C also illustrate that the survival benefit associated with the CC rs419598 genotype diminished as patients approached the age of 80. To further assess this finding we analyzed differences between genotypes and clinical outcomes in patients above and less than or equal to age of 80. Elevations of inflammatory cytokines did not differ between the two age groups (Supplemental Table 4). However, as expected, there was a marked difference in age, sex and BMI adjusted mortality between cohorts of patients above and below the age of 80 (33.0% vs. 11.7%, OR 3.92 (95% CI 3.04 -5.04); < 0.001) (Supplemental Table 4). The higher mortality was associated with significantly increased pre-existing comorbid risk conditions among the older age group, including coronary artery disease, heart failure, chronic lung disease and cancer (Supplemental Table 5).

We next examined the effects of the *IL1RN* rs419598 CC SNV and the CTA haplotype in patients above and below 80 years. The frequency of the CC rs419598 genotype was comparable between those under vs. those over 80 (6.7% vs. 7.2%, p=0.85). For all patients under 80, the *IL1RN* rs419598 CC SNV was associated with significantly lower mortality [6.9% vs.12%; adjusted OR 0.49 (0.23 – 0.94);0.049] in association with significantly lower inflammatory biomarker levels (IL-1β, IL-2, IL-6) and numerically higher IL-1Ra levels than the CT/TT (Table 6). In patients over 80, with multiple co-morbidities, the CC *IL1RN* SNV was associated with numerically decreased inflammatory markers and increased IL-1Ra that did not reach significance, but these effects conferred no protective effect on mortality (30% vs. 33%) (Table 6). For all patients under 80, the CTA haplotype was associated with lower mortality [OR 0.63 (95%CI 0.35 – 1.14); p=0.178] that did not reach statistical significance after adjusting for age, sex, and BMI. However, as noted above, male carriers of the CTA haplotype between the ages of 55-74 did exhibit reduced mortality.

**Table 6:**
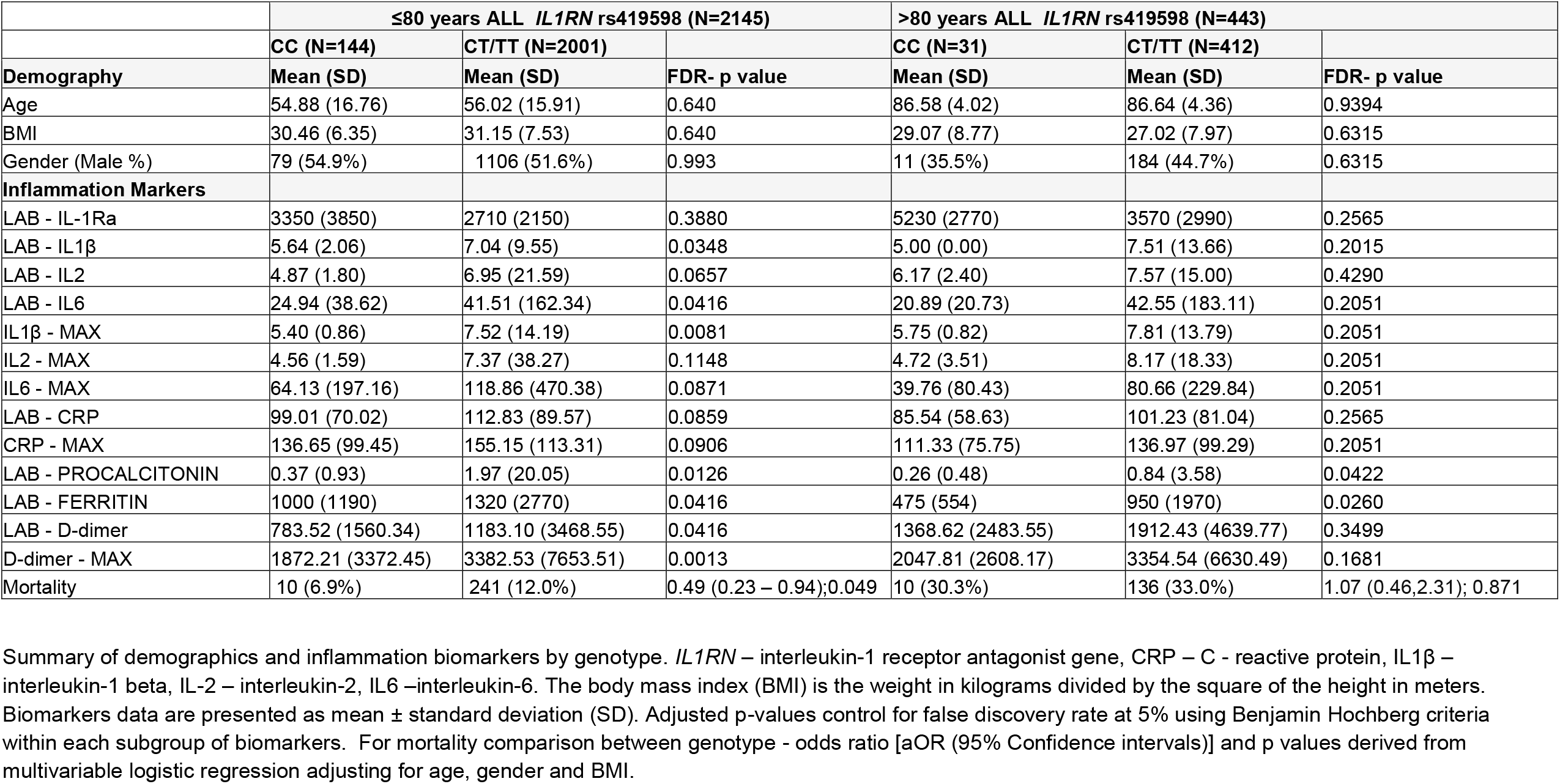
Comparison biomarkers and mortality between ≤80 and >80 years of age according to *IL1RN* rs419598 genotypes.

### Outcomes in Black vs. White Patients

To explore issues related to SARS-CoV-2 outcomes and health equity, we compared self-identified White and Black patients in our population. Table 7 shows that there was no difference in either plasma elevations of inflammatory biomarkers or the mortality between White and Black patients (16.9% vs. 15.7%, p=0.171). Of interest, plasma IL-1Ra levels were lower in Black patients compared to Whites patuents (Table 7) as has been reported (22), and perhaps reflects the differing frequencies of *IL1RN* genotypes. As observed in the total cohort, mortality was greater in Black men than in Black women (19.0% vs. 12.5%, p=0.059). As has been reported, the frequency of the CC rs419598 genotype was lower in Black patients (1.1%) than in White patients (8%), comparable to values reported in the 1000 Genomes Database (23)[Ensembl GRCh37 release 103 - February 2021 © EMBL-EBI]. (Supplemental Table 6). The frequency of one or two copies of the CTA haplotype was 10.2% in White patients and approximately 1% in Black patients. As shown in Supplemental Table 7, CRS and chemical biomarkers of mortality were significantly lower among White patients with the CC rs419598 genotype, and mortality was reduced (10.0% vs. 17.6%, OR 0.54 (0.25 - 1.03); 0.081). Similar trends were observed in White carriers of the CTA haplotype (Supplemental Table 8). Among Black patients, only five individuals carried the CC rs419598 genotype, and no deaths were observed. Given the low numbers of CC carriers, too few laboratory values were available to compare CRS markers between genotypes.

**Table 7.** Association of inflammation markers, and mortality between Black and White Patients.

|  | **Blacks (N=471)** | **Whites (N=1250)** |  |
| --- | --- | --- | --- |
|  | **Mean (SD)** | **Mean (SD)** | **FDR p-value** |
| **Demography** |  |  |  |
| Age | 60.3(17.1) | 64.3(19.6) | < 0.001 |
| BMI | 32.1(8.54) | 30.1(7.97) | < 0.001 |
| Male % | 232 (49.3%) | 643 (51.4%) | 0.4510 |
| **Inflammation Markers** |  |  |  |
| LAB - IL-1Ra | 2250(1410) | 3190(2770) | 0.0007 |
| LAB - IL1β | 8.03(11.4) | 7.57(12.7) | 0.7760 |
| LAB - IL2 | 11.0(44.3) | 6.19(9.74) | 0.5720 |
| LAB - IL6 | 47.1(234) | 37.1(148) | 0.6689 |
| IL1β - MAX | 8.23(12.2) | 7.83(13.3) | 0.7760 |
| IL2 - MAX | 14.3(80.4) | 6.30(11.8) | 0.5720 |
| IL6 - MAX | 110(368) | 92.0(460) | 0.6689 |
| LAB - CRP | 104(81.6) | 108(88.2) | 0.5720 |
| CRP - MAX | 153(114) | 147(109) | 0.5720 |
| LAB - PROCALCITONIN | 5.42(38.8) | 0.844(4.57) | 0.1040 |
| LAB - FERRITIN | 1280(2510) | 1130(2800) | 0.5720 |
| LAB - D-dimer | 1550(4830) | 1190(3020) | 0.5363 |
| D-dimer - MAX | 4650(10400) | 2810(5950) | 0.0007 |
| Mortality % | 74 (15.7%) | 211 (16.9%) | 0.80 (0.59 – 1.10); 0.171 |

Summary of demographics and inflammation biomarkers by race. CRP – C - reactive protein, IL1β – interleukin-1 beta, IL-2 – interleukin 2, IL6 –interleukin-6. The body mass index (BMI) is the weight in kilograms divided by the square of the height in meters. Biomarkers data are presented as mean ± standard deviation (SD). Adjusted p-values control for false discovery rate at 5% using Benjamin Hochberg criteria within each subgroup of biomarkers. For mortality comparison - odds ratio [aOR (95% Confidence intervals)] and p values derived from multivariable logistic regression adjusting for age, gender and BMI.

## Discussion

The results described here are consistent with our prior finding that the CTA *IL1RN* haplotype is associated with decreased clinical severity of both osteoarthritis and rheumatoid arthritis, including lower disease activity scores (DAS), in association with *increased* plasma IL-1Ra and *decreased* plasma IL-6 and CRP levels (19). Genetic polymorphisms of the *IL1RN* gene have also been reported to modulate systemic inflammation in diverse inflammatory syndromes but have not been evaluated in SARS-CoV-2 infection (21, 24, 25). In these studies we utilized low coverage whole genome sequencing of 2589 patients hospitalized patients with SARS-CoV-2 to examine the association of single nucleotide polymorphisms (SNPs) of *IL1RN* with biochemical markers of the cytokine release syndrome and mortality. The novel finding of this report is that a specific haplotype, CTA, and the sequence variant of *IL1RN*, rs419598, are associated with attenuation of the cytokine release syndrome in SARS-CoV-2 and significantly decreased mortality in men.

While much attention has been given to the role of *pro-inflammatory* cytokines that characterize the CRS in acute SARS-CoV-2 infection, there has been less focus on *anti-inflammatory* cytokines. In the current study, we show the anti-inflammatory cytokines, IL-10 and IL-1Ra levels were exceptionally high in hospitalized patients with acute SARS-CoV2 infection. In the case of IL-1Ra, exceeding 3000 pg/ml, eightfold higher than normal values of 200 - 450 pg/ml (https://www.proteinatlas.org/ENSG00000136689-IL1RN/blood+protein), These data are consistent with the findings of Bulow, et.al., that elevated plasma IL-1Ra levels at the time of admission were associated with respiratory failure, acute kidney injury, and mortality in critically ill COVID-19 (26). Moreover, a specific role for IL-1Ra in the regulation of the cytokine release syndrome and clinical severity of COVID-19 severity is supported by the findings of Zhao et al. (11), who showed that IL-1Ra and IL-10, both greater than 10-fold increased in our studies (Figure 1; panel A), were among the four cytokines most predictive of the cytokine release syndrome, with elevations appearing early in patients with severe disease. Zhao et al. speculated that the early elevation of IL-1Ra and IL-10, both “ anti-inflammatory”, is a mechanism to control hyper-inflammation responses during the early stages of immune activation, acting to attenuate the immune system activation and disease severity. Huntington support these findings and utilized a mutual information algorithm that classified the information gained for CRS prediction provided by cytokine expression levels and clinical variables (27). Using this methodology, they found that a small number of cytokine expression variables were associated with COVID-19 disease severity, the most predictive of which included IL-1Ra as well as macrophage colony-stimulating factor (M-CSF), and interferon-inducible protein 10 (IP-10). Recent studies have also highlighted the important role of macrophages in the cytokine storm and systemic inflammation that contributes to COVID-19 pathogenesis (28, 29).

With regard to *IL1RN*, Meyer et al., demonstrated that the C allele of the synonymous *IL1RN* coding variant at rs315952 was associated with reduced risk for acute respiratory distress syndrome (ARDS) and increased plasma IL-1Ra (30). They showed that IL1RN transcripts were differentially expressed depending on which synonymous coding variant was present in the transcript. The variable number of tandem repeat polymorphic at *IL1RN* (24) is in high LD with the two SNVs (rs419598 and rs315952) examined in this study. Taken together, these studies demonstrate that genetic polymorphisms of *IL1RN* influence the severity of diverse inflammatory syndromes, presumably by modulating the levels or function of IL-1Ra, thereby regulating IL-1β driven cascades of pro-inflammatory cytokines and chemokines (25, 31). Previously, Rafiq et al have reported that peripheral blood mononuclear cells from subjects with rs419598 genotype CC had increased IL-1Ra mRNA expression compared with genotype TT (32). Our studies build upon these observations, indicating that the specific sequence variant of *IL1RN*, rs419598, *modulates* the severity of acute SARS-CoV-2 via the regulation of the IL-1Ra response. Finally, Balzanelli et al., recently reported in a small cohort of 41 patients that individuals with the CC rs419598 SNV were less likely to be hospitalized with SARS-CoV2 and exhibited increased plasma levels of IL-1Ra (33).

It is also known that genetic polymorphisms that decrease Type 1 interferon (IFN-I) production and the development of anti-IFN-I autoantibodies are associated with more severe cases of COVID-19 (34, 35). This may be explained, in part, by the lack of IFN responses against SARS-CoV-2 early on during infection (34, 36-38). Our data raise the additional possibility of cross-talk between the IL-1β and Type 1 interferon pathways in the pathogenesis of the cytokine release syndrome in COVID-19. It is noteworthy that in addition to its antiviral properties, IFN-1 is also known to suppress the inflammatory response by inhibiting the inflammasome–IL-1β axis and induce the expression of IL-1Ra (39). Thus, it is possible that both the anti-inflammatory properties of IFN-1 and the IL1RN genetic variation in rs419598 may be mediated by inhibitory effects at the level of the IL-1 β and IFN axis in SARS-CoV-2 infection (39-41).

An intriguing finding of this study is the differential effects of *IL1RN* polymorphisms on mortality between male and female patients. Consistent with prior reports, the mortality of patients hospitalized with COVID-19 was significantly higher in men than women (5). This is notwithstanding our finding that plasma elevations of inflammatory cytokines and d-dimer measured upon admission at baseline were elevated in both men and women. The *IL1RN* rs419598 CC genotype, except for IL-6 Max, which was unaffected by the CC genotype in women, was associated with lower plasma inflammatory markers and higher IL-1Ra levels in both men and women. Interestingly, the levels of IL-1Ra in women with the *IL1RN* rs419598 CC carriers exceeded 4950 pg/ml, two-fold higher than in women with the CT/TT genotype and 18-fold higher than normal. However, while the *IL1RN* CC genotype was associated with lower inflammatory biomarker levels in both sexes, reduced mortality was limited to effects in men in the overall population. We do note that among White women, the trend towards decreased mortality in *IL1RN* CC carriers was observed (8.9% vs. 14.9%, p<0.15). Given that our data were adjusted for age, sex and BMI, further exploration with an increased number of patients stratified by race is warranted in order to reveal race and sex specific effects of *IL1RN* variants Alternatively, other sex-related protective or susceptibility factors may also play a role. There is evidence that IL-1Ra and IL-1 production regulation may be fundamentally different in women than men: for example, plasma IL-1Ra levels have been reported to be higher in men, while the *in vitro* production of IL-1Ra from isolated monocytes is significantly higher in women than in men (17, 42, 43). In addition, androgen receptors, which lead to the activation of promoters for TMPRSS2, can facilitate SARS-CoV-2 entry into cells (44) and may contribute to different outcomes in males versus females. Thus, our data may reflect differential cytokine regulation by the IL-1 pathway and/or androgen effects that merit investigation in the understanding of gender-related outcomes following COVID-19 infection.

There are several limitations to our study. Since we employed low-pass whole genome sequencing for genotype information, coverage may limit ability to detect SNVs and thereby limit interpretations of haplotype and linkage disequilibrium. Since the data were retrieved from the EHR, cytokine data were unavailable for all patients. However, for the key proinflammatory cytokines of CRS (IL-1β, IL-6, IL-2) the number of individual patient values in the EHR were substantial, ranging from 642-1174 (Supplemental Table 1). Moreover, the *IL1RN* SNVs findings regarding inflammatory cytokines parallel the findings for conventional markers of inflammation (CRP, D-dimer, procalcitonin, ferritin), which were available for essentially all patients. We also note that our patient population reflected infection before the Omicron wave and widespread vaccination of the population. However, the clinical presentations and CRS features of Omicron infection in hospitalized patients with acute SARS-CoV-2 infection remain comparable to those observed in prior variants, making it unlikely that the genetic determinants shown here would be substantially different. Finally, whether the *IL1RN* polymorphism effects in COVID-19 or other inflammatory diseases are due to effects on levels of IL-1Ra, or on differential functions of the produced IL-1Ra protein can been speculated, but is not known.

In summary, these studies demonstrate that the *IL1RN* CTA haplotype and rs419598 CC genotype are significantly associated with attenuation of the cytokine release syndrome in patients with severe SARS-CoV2 infection and reduce mortality in men. We show that concomitant with decreased pro-inflammatory cytokine production, the *IL1RN* rs419598 CC genotype is associated with increased levels of its anti-inflammatory gene product IL-1Ra, a measurable biological effect of the distinct polymorphism that we have observed in prior studies. Our data provide genetic evidence that activation of the inflammasome and the IL-1 pathway is proximal in the systemic cytokine inflammatory cascade. Its regulation by IL-1Ra, an endogenous anti-inflammatory protein, and potential cross-talk with IFN require further elucidation to advance the understanding and treatment of SARS-CoV-2 infection.

## Materials and Methods

### Patient population

This is a retrospective, observational cohort of patients admitted to Tisch Hospital, one of three hospitals within the NYU Langone Health (NYULH) system. The Institutional Review Board at NYU Grossman School of Medicine approved the protocol (S16-00122 and i20-00485). Age, sex and BMI matched healthy controls for multiplex cytokines screening were recruited from our Osteoarthritis Clinics who did not have inflammatory co-morbidities and were never exposed to SARS-CoV-2 (#i05-131).

### Study cohort

We identified patients tested for SARS CoV-2 and positive for real-time reverse transcriptase-polymerase chain reaction (RT-PCR) assays on nasopharyngeal or oropharyngeal swab specimens. Our clinical laboratory conducted tests using the Roche SARS-CoV-2 assay in the Cobas 6800 instruments through emergency use authorization granted by the US Food and Drug Administration. On 31 March, we added testing using the SARS-CoV-2 Xpert Xpress assay in the Cepheid GeneXpert instruments under emergency use authorization by the FDA. The targets amplified by these assays are the ORF1/a and E genes in the Roche Cobas assay and N2 and E genes in the Cepheid XpertXpress.

### Data sources

Study data were obtained from the electronic health record (EHR, Epic Systems, Verona, WI), an integrated health record that included all inpatient and outpatient visits in the health system. We obtained demographics, diagnosis codes, procedures, vital signs, and laboratory measurements during hospitalization from these records. Demographic data included age, sex, BMI, self-identified race (White, Black or African-American, Asian, other and unknown), and ethnicity (non-Hispanic/Latino, Hispanic/Latino or unknown). All vital signs and laboratory values were obtained as part of their indicated clinical care. Additionally, we obtained co-morbidities in the EHR at the time of presentation, including the history of hypertension, hyperlipidemia, coronary artery disease, heart failure, pulmonary disease (defined by chronic obstructive pulmonary disease or asthma), malignancy (excluding non-metastatic non-melanoma skin cancer), diabetes, and obesity (defined by their most recent BMI). We evaluated the initial and maximum values during hospitalization for markers of inflammation, including ferritin, interleukin-1β (IL-1β), interleukin-2 (IL-2), interleukin-6 (IL-6), C-reactive protein (CRP), and D-dimer. Variable numbers of specimens for non-routine laboratory tests (e.g., complement C3 and C4) are shown in Supplemental Table 1.

### Sample processing, genotyping, and imputation

Discarded blood samples for clinical use were utilized in this study. Total genomic DNA was isolated using standard protocols and used to generate 1.2X low coverage (lc) human whole-genome sequence (WGS) using standard protocols (20) in the NYU Langone Genome Technology Center. After quality control, these data were used to impute all common (minor allele frequency ≥ 1%) SNV genotypes for each sample, using a reference population of 7,345 samples from 26 distinct geographic populations of the world (gencove.org); these analyses were conducted by Gencove (New York, NY) and have been demonstrated to produce genotype data with non-reference allele concordance ≥ 98% (20). We extracted *IL1RN* (rs419598, rs315952, and rs9005) genotypes for this study; allele frequencies were estimated using standard allele counting. Plasma cytokines IL-1β, IL-2, and IL-6 were determined by a test developed by ARUP Laboratories (Salt Lake City, UT 84108) and approved by the New York State Department of Health).

### Multiplex assay

Plasma samples for the sub-set of SARS-Cov-2 patients’ (n=359) samples and healthy controls (n=22) were screened to measure the levels of multiple cytokines: CCL2, IL-8, IFNα, IFN-β, IFN-γ, IL-10, IL-12p70, IL-13, IL-17A, IL-1β, IL-1RA, IL-2, IL-5, IL-6, TNF-α, VEGF, and IL-1α. A customized version of the Human XL Cytokine Premixed Kit (#FCSTM18, R&D Systems) was used to simultaneously measure the 17 analytes through a magnetic bead–based fluorescence sandwich ELISA. Samples were thawed on ice, centrifuged at 10,000 x g for 10 minutes to remove particulates, and then added to premixed beads, according to the manufacturer’s protocol. A Luminex® 200TM instrument was used to collect and measure the Median Fluorescent Intensity (MFI) of 50 beads per analyte per well, following kit’s instructions. The assay contained recombinant protein standards (Percentage of recovery within 75%-120% range) and quality control (QC) samples (high and low concentration) for each analyte. A 4- or 5-parameter logistic function with 1/y2 weighting was fit using Belysa® v.1 analysis software (MilliporeSigma) to calculate each cytokine’s concentration. The lower limit of quantitation was defined by the software for each analyte.

### Haplotype analysis

All three SNPs (rs419598, rs315952, and rs9005) studied in this study are in the *IL1RN* gene; we evaluated haplotype effects on CRS and mortality as described below. Two of the nine potential haplotypes that could be constructed from these three SNPs occurred with a frequency of >1% (haplotypes CTA and TTG). Both CTA and TTG are found on the same locus. Specifically, 61.7% of subjects could be unambiguously inferred to carry 0, 1, or 2 copies of the TTG haplotype, and 12% of subjects could be unambiguously inferred to have 1 or 2 copies of the CTA haplotype. Throughout this report, we denote the CTA-1/2 or TTG-1 or TTG-2 haplotype groups to represent carriers of 0, 1, or 2 copies of the *IL1RN* TTG haplotypes. The frequencies of the *IL1RN* haplotypes forn NYU COVID-19 cohort of 2589 participants, the frequencies of TTG-0, TTG-1 and TTG-2 were 11.4%, 46.1% and 16.6%, respectively. The overall frequency of the CTA-1 or CTA-2 haplotype was 10.2% (Supplemental Table 6).

### Primary outcomes and study variables

Patient outcomes of interest included inflammatory markers for evidence of cytokine release syndrome (CRS) and mortality as evidence of severe COVID-19 outcome. Other study variables included age, sex, self-identified race and ethnicity, BMI, and pre-existent co-morbidities.

### Statistical analysis

Continuous variables were summarized using means and standard deviations, whereas categorical variables were summarized using frequency and proportions. These summaries were created to assess demographic variables and CRS in the overall study sample and by mortality status (alive and deceased), by sex (men and women) and by race/ethnicity. Next, we assessed demographic variables, CRS and mortality stratified by *IL1RN* genotypes/haplotypes, overall and separately for males and females. Similar summaries were also generated to compare observed differences in Non-Hispanic Black versus Non-Hispanic White patients and among those ≤80 years and >80 years of age. Univariate parametric tests (t-test for continuous variables and *χ*^2^ tests for categorical variables) were used to assess CRS and mortality outcomes by each strata. Separate age, sex and BMI adjusted logistic regressions were fitted to estimate the odds ratios (ORs) and 95% confidence interval (CI) for predicting mortality with each genotype/haplotype as the primary covariate. Significance tests were two-tailed at the significance level of 0.05. To account for multiple comparisons, p-values from univariate tests were adjusted at a 5% false discovery rate using the Benjamin Hochberg criterion, separately for demographics and CRS category. Correlation between elevated levels of interferon, cytokines and chemokines were assess using Spearman correlation test.

For primary analysis to compare in-hospital mortality between *IL1RN* genotypes, we used multivariable logistic regression, adjusting for age, sex, and BMI in a stepwise procedure. Age was adjusted first as a continuous variable. To account for non-linear relationships between age and COVID-19 mortality, we adjusted for categorical age in deciles as a secondary analysis. ORs and 95% CIs were reported. Further, to assess time to in-hospital mortality between genotypes, we used the Cox proportional hazards model, adjusting for age, sex, and BMI. Hazard ratios and 95% confidence intervals were reported. Finally, these analyses were repeated separately for sex-stratified samples to assess differences in relationships between males and females. The statistical software R was used for the statistical analysis.

## Data Availability

All data produced in the present study are available upon reasonable request to the authors

## Author Contributions

MA, CP and SA contributed equally to work. SBA, MA, and AC contributed to study concept and design. MA, CP, SA, EI, XL, ST, AB, NH, AC, and SBA contributed to literature search, writing the manuscript and data interpretation. MA, CP, SA, EI, XL, ST, NH, AC and SBA participated in data collection and data analysis. MA, SA and XL made figures and tables.

## Acknowledgments

SBA was supported by National Institutes of Health R21 - AR078466P. We would like to acknowledge the Immune Monitoring Laboratory (Division of Advanced Research Technologies, DART) at NYU Langone Health, partially supported by NIH grant P30CA016087.

## Notes

### Competing Interest Statement

The authors have declared no competing interest.

### Funding Statement

Steven B. Abramson was partially supported by National Institutes of Health R21 - AR078466P

### Author Declarations

The Institutional Review Board at NYU Grossman School of Medicine approved the protocol (S16-00122, i20-00485 and #i05-131).

