## Supplemental Figures and Tables for "Interleukin-1 receptor antagonist gene (*IL1RN*) variants modulate the cytokine release syndrome and mortality of SARS-CoV-2"

**Supplemental Figure 1: Mortality protected among 55-74 years age groups of IL1RN haplotype CTA-1/2 carriers.**

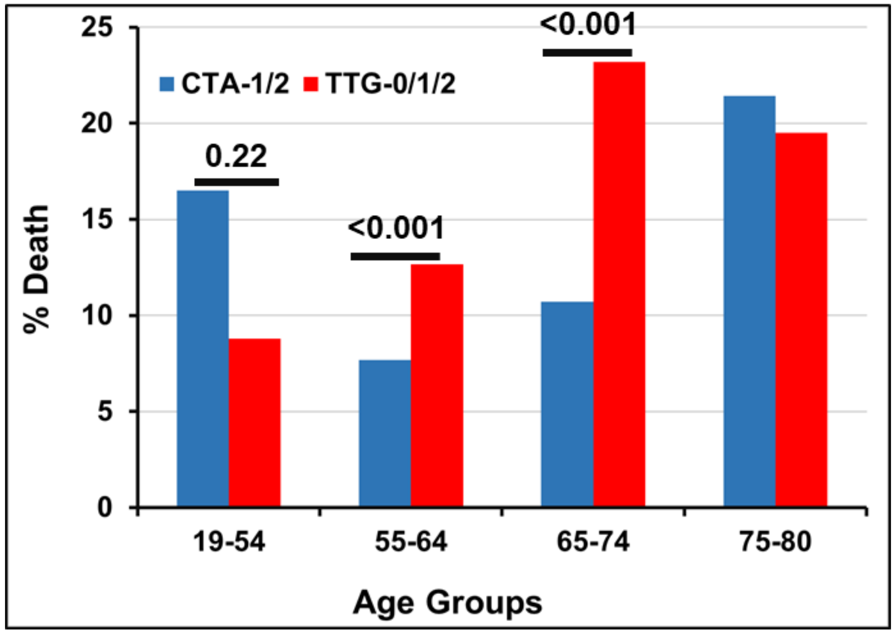


| | **% Mortality Among Men *IL1RN* Haplotypes** | |
|---|---|---|
| **Age (years)** | **CTA-1/2 (N=150)** | **TTG-0/1/2 (N=1008)** |
| 19-44 | 2.9% (35) | 2.9% (136) |
| 45-54 | 13.6% (22) | 5.9% (136) |
| 55-64 | 7.7% (39) | 12.7% (229) |
| 65-74 | 10.7% (28) | 23.2% (276) |
| 75-80 | 21.4% (14) | 19.5% (82) |

**SUPPLEMENTAL FIGURE 1: IL1RN CTA 1/2 Protective Haplotype is Associated with Decreased Mortality Among Men.**
**2A**. Comparison of mortality rate among men between IL1RN CTA-1/2 vs. TTG-0/1/2 haplotype by decade.
**2B.** Mortality as percentage is shown for *IL1RN* CTA and TTG-0/1/2 carriers among men. Total number of patients are shown in parentheses for each age group.

**Supplemental Table 1: Demography, inflammation biomarkers levels in COVID-19 patients.**

| | Number of observations | Plasma Levels Mean (SD) |
|---|---|---|
| **Demography** | | |
| Age | 2589 | 61.2 (18.7) |
| BMI | 2532 | 30.43 (7.71) |
| Sex (% Male) | 1380 (53.3%) | |
| **Inflammation Markers** | | |
| LAB - IL-1Ra | 465 | 2897.28 (2449.21) |
| LAB - IL1β | 642 | 7.02 (10.05) |
| LAB - IL2 | 645 | 6.92 (19.89) |
| LAB - IL6 | 1229 | 40.76 (161.30) |
| IL1β - MAX | 642 | 7.43 (13.64) |
| IL2 - MAX | 645 | 7.31 (34.37) |
| IL6 - MAX | 1174 | 107.10 (421.30) |
| LAB - CRP | 2031 | 109.50 (86.96) |
| CRP - MAX | 1984 | 152.00 (109.60) |
| LAB - PROCALCITONIN | 1928 | 1.68 (17.50) |
| LAB - FERRITIN | 2022 | 1232.0 (2583.0) |
| LAB – D-dimer | 2046 | 1290.0 (3634.0) |
| D-dimer - MAX | 2039 | 3277.0 (7282.0) |
| LAB - C3 COMPLEMENT | 176 | 117.3 (39.82) |
| LAB - C4 COMPLEMENT | 172 | 30.42 (14.91) |
| Mortality | 397 (15.3%) | |

IL-1Ra – interleukin-1 receptor antagonist, CRP – C - reactive protein, IL1β – interleukin-1 beta, IL2 – interleukin-2, IL6 –interleukin-6. The body mass index (BMI) is the weight in kilograms divided by the square of the height in meters. Biomarkers data are presented as mean ± standard deviation (SD).

**Supplemental Table 2: Association of plasma inflammation cytokine biomarkers and mortality - with the *IL1RN* rs315952 *CC* genotype**

| | ***IL1RN rs315952 (N= 2589)*** | | |
|---|---|---|---|
| | **CC (N=237)** | **CT/TT (N=2352)** | |
| **Demography** | **Mean (SD)** | **Mean (SD)** | **FDR- p value** |
| Age | 61.09 (17.39) | 61.21 (18.78) | 0.9190 |
| BMI | 30.13 (7.75) | 30.88 (21.15) | 0.8955 |
| Male % | 90 (51.4%) | 1290 (53.4%) | 0.8955 |
| **Inflammation Markers** | | | |
| IL-1Ra | 2650 (2160) | 2940 (2500) | 0.4893 |
| LAB - IL1 | 5.40 (0.86) | 7.19 (10.54) | 0.0007 |
| LAB - IL2 | 4.84 (2.22) | 7.14 (20.88) | 0.0520 |
| LAB - IL6 | 34.92 (77.32) | 41.15 (167.57) | 0.5168 |
| IL1 - MAX | 5.42 (0.87) | 7.64 (14.32) | 0.0007 |
| IL2 - MAX | 5.15 (4.88) | 7.54 (36.08) | 0.2581 |
| IL6 - MAX | 94.43 (224.73) | 109.47 (438.63) | 0.5500 |
| LAB-CRP | 119.88 (90.64) | 108.65 (86.58) | 0.2581 |
| CRP - MAX | 157.87 (110.06) | 149.62 (110.13) | 0.4839 |
| LAB - PROCALCITONIN | 3.96 (42.07) | 1.41 (12.27) | 0.4893 |
| LAB - FERRITIN | 1599.22 (3561.04) | 1186.84 (2437.36) | 0.2581 |
| LAB – D-dimer | 1932.25 (4818.88) | 1233.02 (3495.84) | 0.1820 |
| D-dimer - MAX | 4048.79 (8573.90) | 3207.75 (7133.68) | 0.3283 |
| Mortality | 40 (16.9%) | 357 (15.2%) | 1.11 (0.74, 1.63); 0.61 |

Summary of demographics and inflammation biomarkers by genotype. CRP – denotes – C - reactive protein, IL1β – interleukin-1 beta, IL-2 – interleukin-2, IL6 – interleukin-6. The body mass index (BMI) is the weight in kilograms divided by the square of the height in meters. Biomarkers are presented as mean ± standard deviation (SD). Adjusted p-values control for false discovery rate at 5% using Benjamin Hochberg criteria within each subgroup of biomarkers.  For mortality comparison between genotypes - odds ratio [aOR (95% Confidence intervals)] and p values derived from multivariable logistic regression adjusting for age, gender and BMI.

**Supplemental Table 3: Association of plasma inflammation cytokine and mortality with the *IL1RN* rs9005 *AA* genotype**

| | ***IL1RN rs9005 (N= 2589)*** | | |
|---|---|---|---|
| | **AA (N=296)** | **AG/GG (N=2293)** | |
| **Demography** | **Mean (SD)** | **Mean (SD)** | **FDR- p value** |
| Age | 57.96 (17.95) | 61.62 (18.71) | 0.0030 |
| BMI | 30.75 (7.20) | 30.40 (7.77) | 0.4420 |
| Male % | 167 (56.4%) | 1213 (52.9%) | 0.4245 |
| **Inflammation Markers** | | | |
| IL-1Ra | 3810 (3640) | 2800 (2270) | 0.1207 |
| LAB - IL1β | 5.80 (2.96) | 7.19 (10.64) | 0.0607 |
| LAB - IL2 | 5.06 (2.10) | 7.17 (21.17) | 0.0715 |
| LAB - IL6 | 52.75 (280.97) | 39.05 (139.55) | 0.7245 |
| IL1β - MAX | 5.84 (2.67) | 7.65 (14.49) | 0.0520 |
| IL2 - MAX | 4.61 (2.21) | 7.68 (36.60) | 0.1062 |
| IL6 - MAX | 119.13 (390.96) | 106.65 (426.85) | 0.7963 |
| LAB - CRP | 108.61 (80.24) | 109.81 (87.82) | 0.8340 |
| CRP - MAX | 139.79 (102.60) | 151.68 (110.97) | 0.1690 |
| LAB - PROCALCITONIN | 0.56 (1.29) | 1.79 (18.46) | 0.0520 |
| LAB - FERRITIN | 1049.35 (1634.20) | 1246.97 (2656.75) | 0.1690 |
| LAB - D-dimer | 1177.93 (3766.41) | 1310.93 (3623.26) | 0.7245 |
| D-dimer - MAX | 2521.78 (5413.44) | 3377.50 (7470.43) | 0.0858 |
| Mortality | Death = 35 (11.7%) | Death = 362 (15.8%) | 0.87 (0.58, 1.27); 0.48 |

Summary of demographics and inflammation biomarkers by genotype. CRP – denotes – C - reactive protein, IL1β – interleukin-1 beta, IL-2 – interleukin-2, IL6 – interleukin-6. The body mass index (BMI) is the weight in kilograms divided by the square of the height in meters. Biomarkers are presented as mean ± standard deviation (SD). Adjusted p-values control for false discovery rate at 5% using Benjamin Hochberg criteria within each subgroup of biomarkers. For mortality comparison between genotypes - odds ratio [aOR (95% Confidence intervals)] and p values derived from multivariable logistic regression adjusting for age, gender and BMI.

**Supplemental Table 4: Comparison inflammation biomarkers between ≤80 and >80 years age group**

| | ALL (N=2589) | | |
|---|---|---|---|
| | ≤80 years (N=2145) | >80 Years (N=443) | |
| **Demography** | **Mean (SD)** | **Mean (SD)** | **FDR – p value** |
| Age | 55.95 (15.96) | 86.63 (4.33) | <0.001 |
| BMI | 31.11 (7.46) | 27.16 (8.04) | <0.001 |
| Gender (male %) | 1185 (55.2%) | 195 (44.0%) | <0.001 |
| **Inflammation Markers** | | | |
| LAB - IL-1Ra | 2751.45 (2313.42) | 3706.57 (2988.51) | 0.0124 |
| LAB - IL1β | 6.94 (9.23) | 7.38 (13.29) | 0.9630 |
| LAB - IL2 | 6.80 (20.82) | 7.49 (14.60) | 0.9630 |
| LAB - IL6 | 40.44 (157.38) | 41.05 (176.82) | 0.9630 |
| IL1β - MAX | 7.37 (13.69) | 7.70 (13.42) | 0.9630 |
| IL2 - MAX | 7.17 (36.89) | 7.99 (17.86) | 0.9630 |
| IL6 - MAX | 115.28 (457.68) | 77.58 (222.28) | 0.1450 |
| LAB - CRP | 111.96 (88.51) | 100.24 (79.84) | 0.0315 |
| CRP - MAX | 154.01 (112.57) | 135.33 (98.08) | 0.0124 |
| LAB - PROCALCITONIN | 1.86 (19.40) | 0.80 (3.47) | 0.1030 |
| LAB – D-dimer | 1156.80 (3377.45) | 1876.81 (4530.08) | 0.0124 |
| D-dimer - MAX | 3286.07 (7462.52) | 3268.96 (6450.67) | 0.9630 |
| rs419598 (CC%) | 144 (6.7%) | 31 (7.2%) | 0.9099 |
| Mortality | 251 (11.7%) | 146 (33.0%) | 3.92 (3.04, 5.04); < 0.001 |

Summary of demographics and inflammation biomarkers by genotype. CRP – denotes – C - reactive protein, IL1β – interleukin-1 beta, IL-2 – interleukin-2, IL6 – interleukin-6. The body mass index (BMI) is the weight in kilograms divided by the square of the height in meters. Biomarkers are presented as mean ± standard deviation (SD). Adjusted p-values control for false discovery rate at 5% using Benjamin Hochberg criteria within each subgroup of biomarkers. For mortality comparison between age groups - odds ratio [aOR (95% Confidence intervals)] and p values derived from multivariable logistic regression adjusting for gender and BMI.

Supplemental Table 5: Co-morbidities and mortality among ≤80 and >80 years age group

| | ≤80 years (N=2145) | >80 Years (N=443) | OR (95% CI); p value |
|---|---|---|---|
| Age (years) | 56.0 (16.0) | 86.6 (4.3) | 0.0001 |
| BMI | 31.1 (7.46) | 27.16 (8.04) | 0.0001 |
| Gender (Male %) | 1185 (55.2%) | 195 (44.1%) | 0.0001 |
| Metabolic Syndrome | 248 (11.6%) | 38 (8.6%) | 1.28 ( 0.86 , 1.86 ) ; 0.212 |
| CV | 1299 (60.6%) | 398 (89.8%) | 7.65 ( 5.51 , 10.88 ) ; < 0.001 |
| Pulm | 345 (16.1%) | 74 (16.7%) | 1.23 ( 0.92 , 1.64 ) ; 0.161 |
| Diabetes | 724 (33.8%) | 145 (32.7%) | 1.21 ( 0.96 , 1.52 ) ; 0.103 |
| Coronary Artery Disease | 252 (11.7%) | 139 (31.4%) | 4.15 ( 3.21 , 5.37 ) ; < 0.001 |
| Heart Failure | 187 (8.7%) | 108 (24.4%) | 4.32 ( 3.25 , 5.75 ) ; < 0.001 |
| Hyperlipdemia | 828 (38.6%) | 285 (64.3%) | 3.49 ( 2.79 , 4.39 ) ; < 0.001 |
| Hypertension | 1075 (50.1%) | 361 (81.5%) | 5.59 ( 4.29 , 7.37 ) ; < 0.001 |
| Peripheral Vascular Disease | 131 (6.10%) | 47 (10.6%) | 2.02 ( 1.4 , 2.87 ) ; < 0.001 |
| Asthma | 259 (12.1%) | 39 (8.8%) | 0.82 ( 0.56 , 1.17 ) ; 0.279 |
| COPD | 130 (6.1%) | 55 (12.4%) | 2.5 ( 1.75 , 3.54 ) ; < 0.001 |
| Dialysis | 108 (5.0%) | 13 (2.9%) | 0.54 ( 0.28 , 0.96 ) ; 0.05 |
| Cancer | 196 (9.1%) | 100 (22.6%) | 2.93 ( 2.21 , 3.88 ) ; < 0.001 |
| CKD | 285 (13.3%) | 68 (15.3%) | 1.32 ( 0.97 , 1.77 ) ; 0.072 |
| Mortality | 251 (11.7%) | 146 (33.0%) | 3.92 (3.04, 5.04); < 0.001 |

Age in years and BMI are presented as mean ± standard deviation (SD). Co-morbidities number of patients reported at admission and (%) over total number of subjects for each group is presented. For age, gender and BMI adjusted p-values control for false discovery rate at 5% using Benjamin Hochberg criteria within each subgroup of biomarkers. For comorbidities comparison between age groups - odds ratio [aOR (95% Confidence intervals)] and p values derived from multivariable logistic regression adjusting for gender and BMI.

**Supplemental Table 6: *IL1RN* rs419598, rs9005 and rs315952 variant genotypes and haplotypes frequency in COVID-19 patients.**

| RACE | Whites (N=1250) | Blacks (N=471) | East Asians (N=105) | South Asians (N=75) | Hispanic (N=419) |
|---|---|---|---|---|---|
| **Total no. of subjects N=2589 (*IL1RN* - rs419598)** | | | | | |
| CC | 100 | 5 | - | 7 | 36 |
| CT | 504 | 65 | 16 | 21 | 174 |
| TT | 646 | 401 | 89 | 47 | 209 |
| CC Genotype Frequency (**%**) NYU- COVID-19 | **8.0** | **1.1** | **0.0** | **9.3** | **8.6** |
| 1000 Genomes Project **CC** genotype Frequency (**%**) | 8.2 | 0.2 | 0.6 | 9.2 | 11.8 |
| **(*IL1RN* – rs315952)** | | | | | |
| CC | 68 | 93 | 28 | 7 | 25 |
| CT | 460 | 230 | 52 | 19 | 180 |
| TT | 722 | 148 | 25 | 49 | 214 |
| CC Genotype Frequency (**%**) NYU- COVID-19 | **5.4** | **19.7** | **26.7** | **9.3** | **6.0** |
| 1000 Genomes Project **CC** genotype Frequency (**%**) | 9.9 | 21.2 | 30.6 | 2.9 | 4.9 |
| **(*IL1RN* - rs9005)** | | | | | |
| AA | 138 | 14 | 10 | 5 | 85 |
| AG | 542 | 152 | 50 | 37 | 198 |
| GG | 570 | 305 | 45 | 33 | 136 |
| AA Genotype Frequency (**%**) NYU- COVID-19 | **11.0** | **2.96** | **9.5** | **6.7** | **20.3** |
| 1000 Genomes Project **AA** genotype Frequency (%) | 8.7 | 2.9 | 10.9 | 15.7 | 26.8 |
| ***IL1RN* haplotype (rs419598, rs315952 and rs9005)** | | | | | |
| CTA-1 | 56 (4.5%) | 2 (0.4%) | 5 (4.8%) | 3 | 38 (9.1%) |
| CTA-2 | 80 (6.4%) | 3 (0.6%) | 0 (0.0%) | 4 | 29 (6.9%) |
| TTG-1 | 523 (41.8%) | 251 (53.3%) | 61 (58.1%) | 32 | 204 (48.7%) |
| TTG-2 | 246 (19.7%) | 90 (19.2%) | 2 (1.9%0 | 16 | 35 (8.4%) |
| CTA-1/TTG-1 | 257 (20.6%) | 27 (5.7%) | 3 (2.9%) | 18 | 64 (15.3%) |
| CTA-0/TTG-0 | 88 (7.0%) | 98 (20.8%) | 33 (31.4%) | 2 | 49 (11.7%) |

**Supplemental Table 7. Comparison of biomarkers - IL1RN - rs419598 CC vs. CT/TT – Whites only**

| | IL1RN rs419598 (N=1250) | | |
|---|---|---|---|
| | **CC (N=100)** | **CT/TT (N=1150)** | |
| **Demography** | **Mean (SD)** | **Mean (SD)** | **FDR – p value** |
| Age | 62.95 (20.73) | 64.46 (19.50) | 0.4830 |
| BMI | 30.66 (7.55) | 30.07 (8.00) | 0.4830 |
| Male % | 44 (44.0%) | 599 (52.1%) | 0.4440 |
| **Inflammation Markers** | | | |
| LAB - IL-1Ra | 3790 (4010) | 3130 (2620) | 0.4670 |
| LAB - IL1β | 5.40 (0.89) | 7.72 (13.16) | 0.0153 |
| LAB - IL2 | 4.42 (1.19) | 6.32 (10.07) | 0.0153 |
| LAB - IL6 | 22.57 (24.88) | 38.21 (153.85) | 0.0579 |
| IL1β - MAX | 5.47 (0.92) | 8.00 (13.76) | 0.0153 |
| IL2 - MAX | 4.28 (1.29) | 6.45 (12.20) | 0.0160 |
| IL6 - MAX | 37.32 (69.39) | 96.27 (476.65) | 0.0295 |
| LAB - CRP | 89.76 (70.45) | 109.66 (89.34) | 0.0416 |
| CRP - MAX | 121.70 (96.40) | 149.32 (110.05) | 0.0416 |
| LAB - PROCALCITONIN | 0.50 (1.12) | 0.87 (4.74) | 0.0826 |
| LAB - FERRITIN | 827.15 (1115.86) | 1155.42 (2896.63) | 0.0579 |
| LAB - D-dimer | 827.82 (1637.43) | 1223.58 (3107.91) | 0.0798 |
| D-dimer - MAX | 1629.51 (2703.57) | 2908.58 (6138.77) | 0.0013 |
| Mortality | 10 (10.0%) | 201 (17.6%) | 0.54 (0.25 - 1.03); 0.081 |
| Mortality - Men only | 5 (11.4%) | 120 (20.0%) | 0.44 (0.14 - 1.08); 0.099 |
| Mortality - Women only | 5 (8.9%) | 81 (14.7%) | 0.70 (0.23 - 1.74); 0.478 |

Summary of demographics and inflammation biomarkers by genotype. *IL1RN* – interleukin-1 receptor antagonist gene, CRP – C - reactive protein, IL1β – interleukin-1 beta, IL-2 – interleukin-2, IL6 –interleukin-6. The body mass index (BMI) is the weight in kilograms divided by the square of the height in meters. Biomarkers data are presented as mean ± standard deviation (SD). Adjusted p-values control for false discovery rate at 5% using Benjamin Hochberg criteria within each subgroup of biomarkers. For overall mortality comparison between genotype - odds ratio [aOR (95% Confidence intervals)] and p values derived from multivariable logistic regression adjusting for age, gender and BMI ; and for age and BMI for gender specific mortality.

**Supplemental Table 8: White patients with the *IL1RN* CTA haplotype carriers exhibit lower plasma inflammation cytokine levels and mortality.**

| | **IL1RN Whites Only (N=1250)** | | | | | |
|---|---|---|---|---|---|---|
| | **CTA-1/2 (N=137)** | **TTG-(0/1/2) (N=856)** | **CTA-1/TTG-1 (N=257)** | **CTA-1/2 Vs. TTG-0/1/2** | **TTG-0/1/2 vs. CTA 1/TTG1** | **CTA-1/2 vs. CTA-1/TTG-1** |
| **Demography** | **Mean (SD)** | **Mean (SD)** | **Mean (SD)** | **FDR - p value** | **FDR - p value** | **FDR - p value** |
| Age | 61.75 (19.18) | 64.60 (19.59) | 64.86 (19.76) | 0.3270 | 0.8550 | 0.3930 |
| BMI | 30.35 (6.64) | 29.91 (7.40) | 30.67 (10.10) | 0.4860 | 0.8070 | 0.7830 |
| Gender (Male %) | 66 (48.2%) | 448 (52.3%) | 129 (50.2%) | 0.4860 | 0.8550 | 0.7830 |
| **Inflammation Markers** | | | | | | |
| IL-1Ra | 4440 (4090) | 2840 (2120) | 4120 (3990) | 0.1011 | 0.4977 | 0.9380 |
| LAB - IL1β | 6.39 (4.05) | 8.04 (14.87) | 6.52 (5.19) | 0.2069 | 0.4977 | 0.9380 |
| LAB - IL2 | 4.93 (2.77) | 6.43 (10.91) | 6.03 (7.31) | 0.1413 | 0.7450 | 0.6419 |
| LAB - IL6 | 28.51 (51.58) | 40.54 (170.09) | 29.33 (85.70) | 0.2640 | 0.5558 | 0.9380 |
| IL1β - MAX | 6.58 (4.01) | 8.31 (15.52) | 6.81 (5.69) | 0.2069 | 0.4977 | 0.9380 |
| IL2 - MAX | 4.57 (2.93) | 6.33 (11.22) | 7.16 (15.95) | 0.1011 | 0.7450 | 0.6419 |
| IL6 - MAX | 49.81 (83.69) | 105.95 (546.06) | 66.92 (134.65) | 0.1011 | 0.4977 | 0.6419 |
| LAB - CRP | 94.68 (74.69) | 111.59 (89.26) | 103.44 (90.08) | 0.1011 | 0.4977 | 0.6419 |
| CRP - MAX | 123.93 (94.84) | 151.71 (108.40) | 144.24 (117.38) | 0.0338 | 0.6175 | 0.6419 |
| LAB - PROCALCITONIN | 0.59 (1.12) | 0.92 (5.30) | 0.72 (2.34) | 0.2069 | 0.6175 | 0.7323 |
| LAB - FERRITIN | 854.16 (1076.04) | 1192.58 (2918.45) | 1060.03 (2995.67) | 0.0978 | 0.6949 | 0.6419 |
| LAB - D-dimer | 759.87 (1434.46) | 1309.51 (3170.48) | 1023.45 (3077.92) | 0.0228 | 0.4977 | 0.6419 |
| D-dimer - MAX | 1681.82 (3178.80) | 3076.46 (6492.51) | 2491.68 (4971.28) | 0.0013 | 0.4977 | 0.6419 |
| Mortality (ALL) | 15 (10.9%) | 155 (18.1%) | 41 (15.9%) | 0.66 (0.35 - 1.15); 0.163 | 1.18 (0.80 - 1.77); 0.415 | 0.78 (0.39 - 1.50); 0.469 |
| Mortality (Men only) | 7 (10.6%) | 93 (20.7%) | 25 (19.3%) | 0.51 (0.20 - 1.12); 0.117 | 1.13 (0.68 - 1.93); 0.633 | 0.61 (0.21 - 1.58); 0.327 |
| Mortality (women only) | 8 (11.3%) | 62 (15.2%) | 16 (12.5%) | 0.87 (0.36 - 1.92); 0.751 | 1.28 (0.70 - 2.46); 0.443 | 1.20 (0.43 - 3.17); 0.717 |

Summary of demographics and inflammation biomarkers by haplotypes. Biomarkers are presented as mean ± standard deviation (SD).
CRP – denotes – C - reactive protein, IL1β – interleukin-1 beta, IL2- interleukin-2, IL6 – interleukin-6. The body mass index (BMI) is the weight in kilograms divided by the square of the height in meters. FDR p-values control for false discovery rate at 5% using Benjamin Hochberg criteria within each subgroup of biomarkers.
For mortality comparison between haplotypes - odds ratio [aOR (95% Confidence intervals)] and p values derived from multivariable logistic regression adjusting for age, gender and BMI; and for age and BMI for gender specific mortality.
